# Effect of household water treatment and hygiene promotion integration on outpatient treatment for severe acute malnutrition in Senegal: the TISA cluster-randomised controlled trial

**DOI:** 10.64898/2026.07.22.26358662

**Authors:** O Cumming, L Braun, F Siroma Nse’e Nomsili, M Ba, AEB Cabo, J Wells, K Gallandat, L Grignard, A Myers, D Leger, AB Traore, D N’Diaye, S Frison, C MacLeod, M Lê-Lacanette, B Wassonguema, J Lapègue, J Knee, Y Gnokane, A Devort, M Seye, C Opondo, AV Brizuela, DS N’Diaye

## Abstract

**Background:** Severe acute malnutrition (SAM) affects approximately 12 million children globally. The aim of this study was to assess the effectiveness of integrating a household water treatment and hygiene promotion intervention within the national protocol of Senegal for outpatient treatment of children aged 6-59 months of age with uncomplicated SAM.

**Methods:** This was a pragmatic cluster-randomised controlled trial with health centres allocated 1:1 to receive either the standard protocol alone or the standard protocol with the addition of the water treatment and hygiene intervention. The primary outcome of the trial was SAM recovery defined under the national protocol in Senegal, and the secondary outcomes were weight gain, referral to inpatient care, enteric pathogen detection, diarrhoea, and all-cause mortality. Participants were followed up over eight weeks.

**Findings:** Between 22 December 2021 and 20 February 2023, 2,411 children were enrolled in the study with 777 defaulting outpatient treatment during eight weeks’ follow-up. There was no difference between control and intervention groups in the proportion of children recovering from SAM eight weeks post admission (39.8% vs 41.2%; adjusted odds ratio [aOR]: 0.97, 95% Confidence Interval [CI]: 0.66, 1.48). There was no difference in weight gain, referral, enteric pathogen detection or mortality between groups. The prevalence of diarrhoea at eight weeks follow-up was higher in the control group (20.0%) than the intervention group (12.5%), with evidence for a large difference between arms, accounting for between-group differences at baseline (aOR: 0.36; 95%CI: 0.26, 0.50).

**Interpretation:** The integration of household water treatment and hygiene promotion in the standard national protocol did not improve SAM recovery nor the related outcomes of weight gain, referral, enteric pathogen detection or mortality but did reduce diarrhoea. Our results suggest that the integration of this intervention to the standard protocol would not improve SAM outcomes in this setting but would potentially reduce the burden of diarrhoea among this vulnerable group.

**Funding:** This study was supported by the Bureau of Humanitarian Assistance of the United States Agency for International Development and the ACF Foundation.

## Introduction

Severe acute malnutrition (SAM) - or severe wasting and/or nutritional oedema - results from inadequate nutrient intake and/or recurrent illnesses and persists as an issue of global health concern (1). In children, SAM is defined as a weight-for-height z-score (WHZ) that is more than three standard deviations below the median of World Health Organisation (WHO) child growth reference standards and/or nutritional oedema (2). This condition leads to increased susceptibility to infectious diseases, such as diarrhoea and pneumonia, and a high risk of mortality (3). In 2024, 43 million children were affected by wasting globally, of whom 12 million experienced SAM (1).

In its recently updated guidelines, the WHO continues to recommend Community-based Management of Acute Malnutrition (CMAM) for treatment of uncomplicated SAM cases (2). Under this approach, adopted by numerous governments and international agencies, children are treated at home with regular visits to outpatient facilities (2). The CMAM approach is regarded as a successful innovation that has reduced costs to both the health system and the individual, and improved the cost-effectiveness of treatment (4). Despite this success, recovery, referral and relapse rates for CMAM programmes vary greatly between settings, and often fall short of the global standard of 75% recovery (5). Identifying the causes of this variability and developing strategies to mitigate poor treatment outcomes can contribute to more effective programmes.

One factor which may contribute to poor recovery rates in CMAM programmes is environmental health risks at the household level, such as unsafe drinking water, limited or absent hygiene practices and limited access to safe sanitation. Whilst the shift to an outpatient-based model of treatment offers many advantages and has demonstrated improved health outcomes, moving from a more controlled hospital setting to generally less controlled household settings may increase the risk of infection for children at a time when they are particularly susceptible. Water, sanitation and hygiene (WASH) conditions at the household and community level are associated with increased risk of SAM (6), a higher risk of infection during outpatient-treatment (6), longer duration of outpatient-treatment (6), and higher risk of relapse following discharge from outpatient-treatment (7). Furthermore, trials in Pakistan (8) and in Chad (9) reported that the integration of drinking water treatment alone and combined with soap and hygiene promotion respectively resulted in improved recovery outcomes when integrated into CMAM programmes.

In response to this emerging evidence, various international agencies, including WHO (10), UNICEF (11), Action Against Hunger (12) and the World Bank (13), have called for the integration of WASH interventions in strategies to prevent and treat acute malnutrition. In the Sahel region, where there is a high burden of SAM, there have been efforts to integrate WASH interventions within CMAM programmes, and several international agencies have advocated for the provision of a “WASH kit” to children on admission to CMAM treatment programmes to reduce risk of infection and thereby support recovery and prevent complications requiring referral. that the evidence for the effectiveness of integrating water, sanitation and hygiene inventions in out-patient treatment programmes was limited and that further high-quality studies are needed (14). The aim of the “Traitement Intégré de la Sous-Nutrition” (TISA) trial was to assess whether the integration of a household water treatment and hygiene promotion intervention within the national CMAM protocol of Senegal improved recovery among uncomplicated SAM cases receiving outpatient treatment.

## Methods

### Study design and setting

This was a pragmatic trial that sought to evaluate whether this intervention could work under real-world conditions rather than an explanatory trial asking whether the intervention could work under ideal conditions (15). As previously described, the trial was implemented within the national health system, in partnership with the MoH and with the active involvement of nurses responsible for the outpatient treatment of SAM at the health posts (16). A Theory of Change describing how the intervention was hypothesised to change the outcomes of interest, along with the underlying assumptions, guided the design of the trial (Figure 1). This study was a cluster-randomised controlled trial (cRCT) with 86 health centres serving as clusters allocated 1:1 to the intervention or control group (Figure 2). The health centres have responsibility under the Ministry of Health’s national protocol of management of acute malnutrition (Protocole de prise en charge de la malnutrition [PECMA]) for managing uncomplicated SAM cases – that is cases without clinical symptoms of medical complications and who pass an appetite test (17) - and referral of complicated cases for inpatient treatment in hospitals. These centres – or Unité de Récupération et d’Education Nutritionnelle Ambulatoire (UREN) – are charged with the identification of SAM cases and subsequent outpatient treatment through weekly visits to the UREN until discharge.

**Figure 1:**
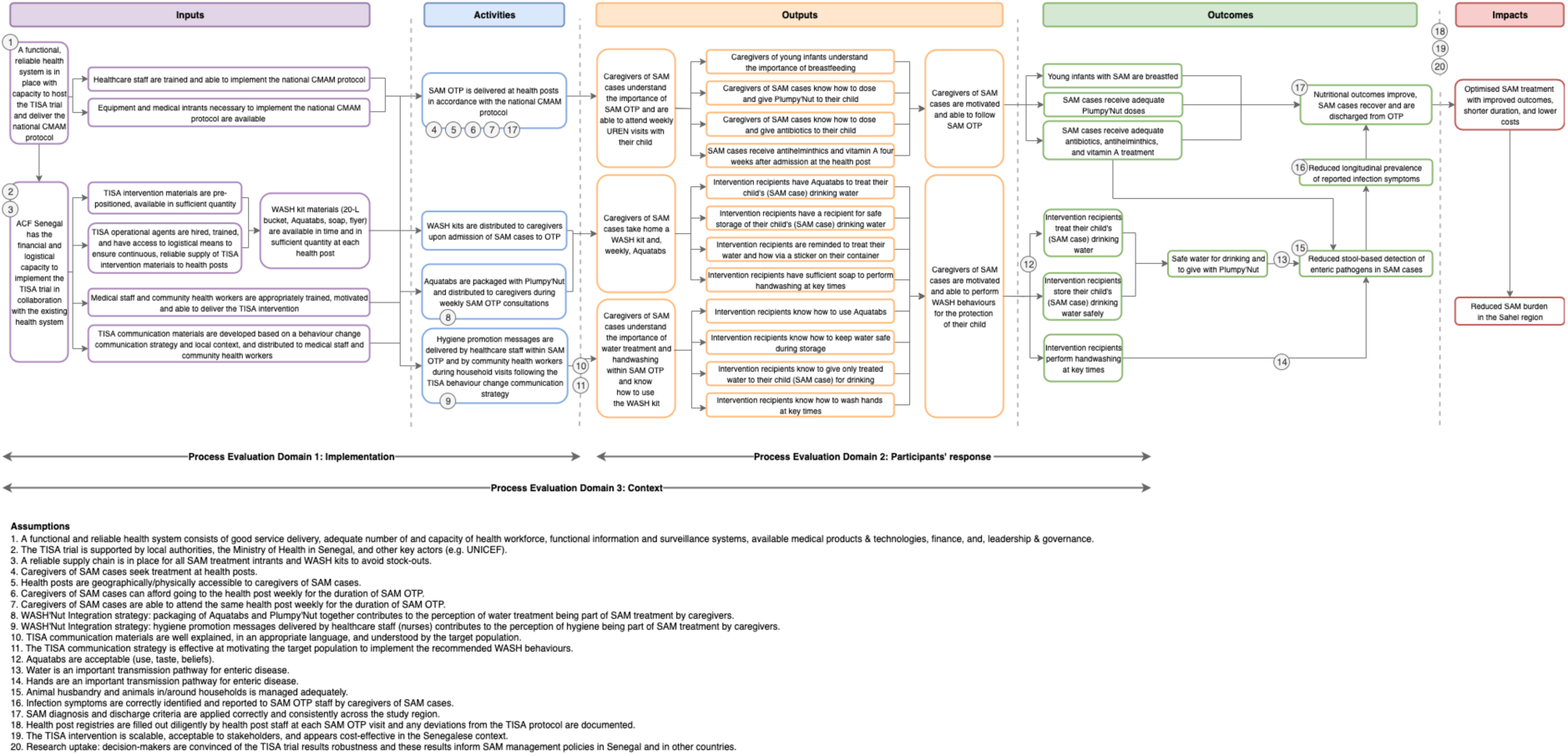
Theory of change for the TISA trial.

**Figure 2:**
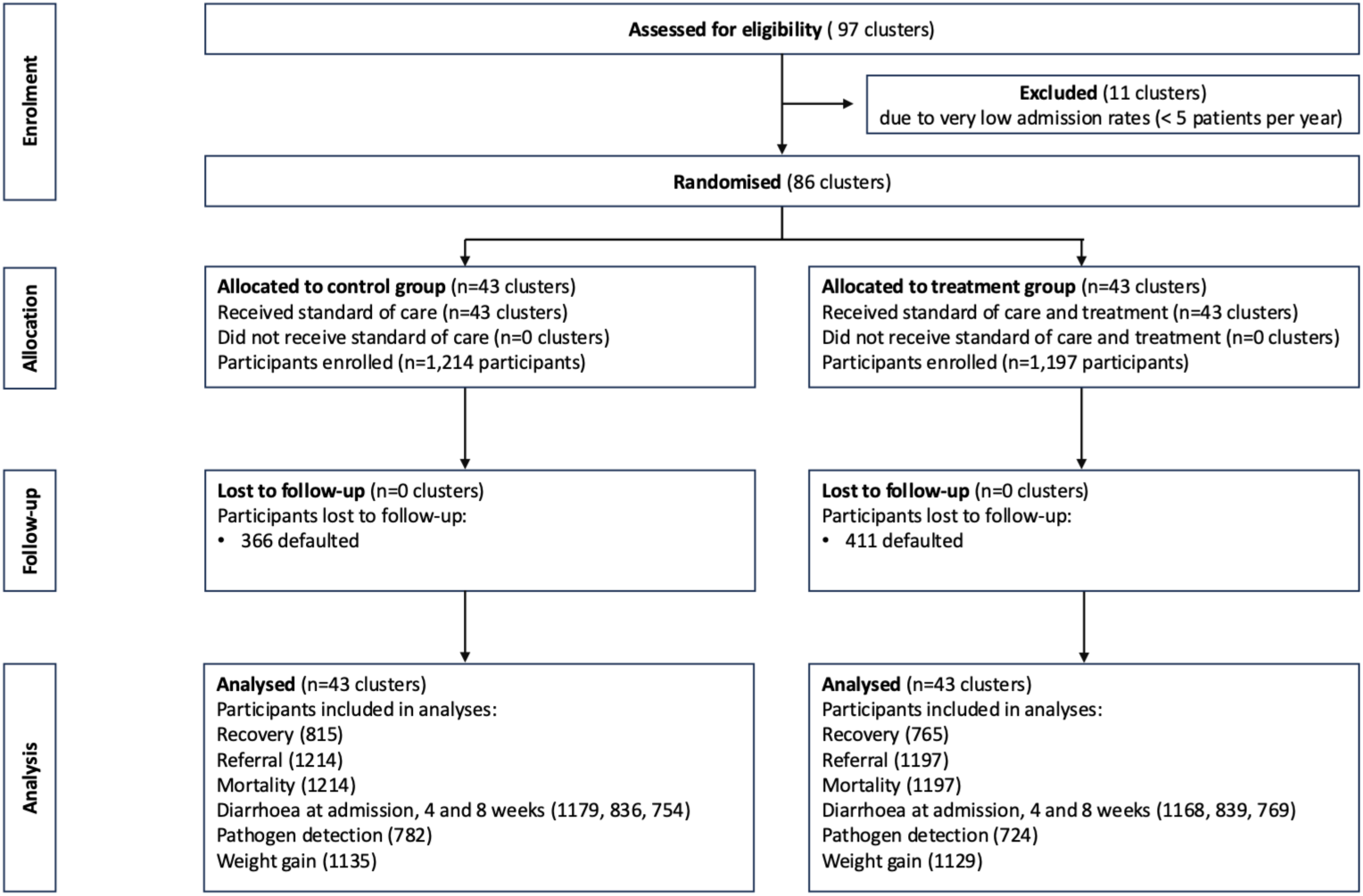
CONSORT study flow diagram.

The trial was conducted across four districts (Podor, Pété, Linguère, and Dahra) of two departments (Podor and Linguère) in northern Senegal. The Government of Senegal identifies these departments as having the highest burden of acute malnutrition and being the highest priority for action on SAM (18). These four districts have a total population of approximately 600,000 and the geographic area covered by the 86 health centres which participated in the trial is approximately 28,000 km^2^. This region has poor access to basic services (health, education and sanitation) and subsistence is based on agriculture or nomadic livestock practices, with traditional labor-intensive practices still prevalent. The Ministry of Health for Senegal aproved the study and facilitated the engagement of the health system through the establishment of local steering committees chaired by the District Medical Officers (“Médecin Chef du District”) for the study sites.

The trial was pre-registered with the ClinicalTrials.gov public registry on 16 December 2020 (NCT04667767). The study protocol was approved by the Comité National d’Ethique pour la Recherche en Santé de Sénégal (000179/MSAS/DPRS/CNERS) and the Research Ethics Committee of the London School of Hygiene and Tropical Medicine in the United Kingdom (17511).

### Participants

All uncomplicated SAM cases among children aged 6-59 months diagnosed in the included health centre were eligible for participation. Inclusion criteria for the study correspond to the diagnostic criteria for SAM used in Senegal: WHZ <-3; or brachial perimeter (mid-upper arm circumference (MUAC)) <115 mm; or bilateral oedema. Patients meeting the diagnostic criteria for SAM, but who also had medical complications are referred to inpatient care and were therefore excluded. Eligibility was assessed by the attending nurse who explained the study to the accompanying caregiver and invited them to participate. All eligible children for whom the parent provided informed consent and confirmed that they intended to remain within the area for the subsequent eight weeks, and who did not meet the exclusion criteria, were enrolled.

Written informed consent was obtained by the senior nurse (ICP; Infirmier Chef de Poste (ICP) at health centers. The ICP explained the study to the caregiver using a standard Participant Information Sheet (PIS) and Consent Statement that was read in Wolof or Pular according to the preference of the caregiver. Hard copies of both documents were provided to all participant caregivers. Only eligible children for whom the parent signed and dated a consent form were enrolled in the study. In the case of illiterate participants, an independent witness signed and dated the consent form, and the participant drew a cross mark in the space provided. Caregivers were informed that they could withdraw from the study any time after enrolment. Any data already collected and analysed was used, unless the participant requested otherwise, but no further analysis was done nor samples kept. No incentives to participate were provided; however, any travel costs to the health centre that would not otherwise be incurred were reimbursed.

### Randomisation and masking

Under the cRCT design, clusters representing the catchment areas for primary health care centres (UREN) were allocated on a one-to-one basis to either a control group receiving the standard outpatient treatment programme (OTP) as per the national protocol for Senegal or an intervention group receiving the OTP plus the household water treatment and hygiene promotion intervention. Clusters were randomly allocated by a statistician at the London School of Hygiene and Tropical Medicine (LSHTM) using a random number generator. This was a public health intervention seeking to change specific behaviours through direct engagement with participants such that blinding of participants to their allocation was not possible nor was it deemed possible to blind the data collection team. Randomisation was conducted remotely but the study investigators and trial statistician were blinded to allocation. The trial statistician conducted the final analyses blinded to allocation.

### Procedures

Participants in both the control and intervention groups received the standard national protocol for outpatient treatment for uncomplicated SAM cases (“la prise en charge de la malnutrition aigüe” [PECMA]) (19). The PECMA is aligned with the WHO Guidelines for the Management and Prevention of Acute Malnutrition (2). Here we briefly describe the standard national protocol for treatment of SAM and then the water treatment and hygiene promotion intervention, the integration of which is the focus of this trial.

The control group received the standard PECMA protocol for outpatient treatment comprising three broad elements. First, medical treatment, whereby patients attend weekly visits at the health centre for anthropometric assessment and appetite testing, antibiotic and anthelminthic treatment, and administration of vitamin A. Second, nutrition treatment whereby the caretaker weekly receives Ready-to-Use Therapeutic Food (RUTF) for the patient based on the weight of the child (measured weekly) together with instruction on how to prepare, store, and administer the RUTF to the child. Third, weekly infant and young child feeding guidance, adapted to the individual child and caregiver.

The intervention group received a water treatment and hygiene promotion intervention integrated with the standard PECMA protocol. This intervention was designed to improve the quality of drinking water consumed by the child and to improve hand hygiene practices around the child whilst they received treatment for SAM. The health promotion component had three key themes: the link between hygiene, infection prevention, and child nutrition; the importance of consistently treating and storing drinking water safely; and the practise of handwashing with soap at five key moments. On admission to treatment, caregivers received a kit comprising a sealable 20 litre (L) bucket with tap and a weekly supply of sodium dichloroisocyanurate tablets (Medentech, Wexford, Ireland), sufficient to treat 20 L/day for seven days. At each subsequent weekly health centre visit, caregivers were provided with a 10-tablet pack alongside RUTF supply. Where attendance the following week was uncertain, nurses provided a two-week supply of both chlorine tablets and RUTF.

Upon admission, caregivers also received two bars of soap and were provided with standardised instruction by the attending nurse on safe hygiene practices, including handwashing after defecation, before preparing food, and before feeding the child, supported by illustrated reference sheets in both Wolof and Pular. The intervention was further reinforced during the weekly visits of the caregiver and child to the health centre by the attending nurse as well as two visits to the household during the period of treatment by a community health volunteer (“Relais Communautaire”).

### Outcomes

The primary outcome of the trial was recovery within eight weeks of admission to outpatient treatment for SAM. Recovery was defined as per the national protocol of Senegal, and in line with the latest WHO Guidelines for Prevention and Management of Wasting (2): two consecutive measures at weekly health centre visits with WHZ ≥ -1.5, if admitted based on WHZ, and/or MUAC ≥ 125 mm, if admitted based on MUAC, and no nutritional oedema. The MUAC and WHZ score, as recorded in the health registry, were used to determine the criteria for admission (i.e., MUAC, WHZ, or both) and recovery was determined based on MUAC (the value recorded in the health registry) and/or the calculated WHZ score using weight and height values recorded in the health registry and computed using the Zscorer R package (20) based on WHO Child Growth Standards (21). Additionally, we conducted an exploratory analysis based on participants’ treatment outcomes as recorded in the health registry by nurses.

An additional five secondary outcomes were all assessed in the eight weeks subsequent to admission to outpatient treatment for SAM. First, weight gain, defined as grams of weight gained by child per kilo per day between admission and exit. Second, referral, defined as the transfer of a participant to hospital due to complications. Third, diarrhoea, defined as occurrence of diarrhoea (three or more loose or liquid stools passed within 24 hours (WHO)) at four and eight weeks (+/-one week) post-admission as reported by the participant’s caregiver in the household questionnaire. Fourth, all-cause mortality defined as any participant death recorded at the health centre during follow-up. And fifth, enteric pathogen detection defined as the stool-based detection of one or more enteric pathogens at eight weeks (+/- one week) post-admission.

Health registry data were recorded by attending nurses at admission and all subsequent weeks of treatment until recovery, discharge, default, referral, transfer or death. Enumerators manually entered the paper health registry records into Open Data Kit for digitisation. Data entry was verified by a remote team against anonymised images of registries to check for consistency and correct any discrepancies. Additionally, household questionnaires were administered during household visits at week 4 and 8 (+/- 1 week) to collect data on WASH kit presence, WASH conditions and practices, and caregiver-reported child and caregiver diarrhoea.

At week 8, nurses collected a rectal swab from participants. Flocked nylon rectal swabs were eluted in 1 mL of liquid Amies solution (eSwab, Copan Diagnostics, catalogue #484CE). Approximately 40 μL of eluate was added to each of four spots on an FTA micro elute card (Qiagen, catalogue # WB120410) and air dried for a minimum of three hours before storage in individual plastic bags with desiccant. Samples were stored in the dark at ambient temperature until shipment to LSHTM for extraction and molecular analysis. We analysed rectal swabs for 30 enteric pathogens (SI Table 4; Supplemental Information (SI)) using a microfluidic qPCR array card (TaqMan Array Card); the laboratory methods have been described previously by our team (22).

### Statistical analysis

All statistical analyses were carried out in Stata version 18 (Stata Corp, College Station, TX) (23). For the original sample size calculation, we used the Hayes-Bennet formula for cRCTs (24). Assuming an intra-class correlation co-efficient of 0.09, and a recovery rate of 67%, based on historical data from the study area, we estimated that with 80% power, and an alpha error of 5% a sample size of 1,720 children across 86 clusters would be sufficient for a minimum detectable difference (MDD) of 10% in the proportion recovered between arms. This MDD was judged appropriate based on the findings of an earlier trial of a very similar intervention (9).

Analysis for all primary and all secondary outcomes was carried out at the individual level with adjustment for clustering within health centres. We adopted an “intention to treat” approach whereby data were analysed according to their allocation to either the intervention or control group, irrespective of participant response to the intervention. Child’s age and gender were adjusted for a priori, and further individual-level variables were adjusted for if they appeared imbalanced between the groups.

For the primary outcome of recovery, we reported the counts and proportions of children recovering in each arm. We also report the odds ratio for recovery in the intervention group relative to the control group with 95% confidence intervals estimated using a mixed effects logistic regression model with random effects at the health centre level to account for clustering. The model was adjusted for age, sex, and variables indicating imbalance at baseline, namely diarrhoea and HAZ at admission, and household sanitation, water source. For weight gain, a mixed effects linear regression model was used to estimate mean differences between groups. For mortality, referral and presence of enteric pathogens, we report the counts and proportions of children in each arm. The odds ratios with 95% confidence intervals were estimated using mixed effects logistic regression models. For the prevalence of diarrhoea, a mixed effects logistic regression model of diarrhoea at four- and eight- weeks post admission adjusted for differences between arms in the prevalence of diarrhoea at admission was used to estimate odds ratios with 95% confidence intervals for the effect of the intervention.

## Results

Between 22 December 2021 and 20 February 2023, 2,411 children diagnosed with uncomplicated SAM and admitted to outpatient treatment were enrolled in the study (Figure 2). The monthly rate of enrolment was similar between arms. 1,214 and 1,197 participants were enrolled in the 43 control and 43 intervention clusters, respectively. Of the enrolled participants, 777 (32.2%) participants – comprising 366 (30.2%) in the control group and 411 (34.3%) in the intervention group – defaulted during treatment before discharge, referral or death. The start of the trial was delayed due to the COVID- 19 pandemic and enrolment was prolonged due to a series of external factors that have been previously described but notably included flooding and national strike action by health workers which affected facilities in the study area as previously described (16).

A greater proportion of participants were male overall (60.0%) and within both the control (60.6%) and intervention (59.3%) arms (Table 1). The mean age of participants at admission was 18.2 months and similar in both arms. Overall, 63.0% of participants were breastfed at the time of admission, and this was similar across arms. Most caregivers were female (94.4%), with a slightly lower proportion in the intervention (92.7%) than the control (96.1%) arm, and a slightly higher proportion of caregivers had not completed primary education in the intervention (73.5%) than the control (69.7%) arm. At admission, the health status of participants was generally less favourable in the intervention arm compared to the control arm. This was most pronounced for child diarrhoea with 44.2% of caregivers in the intervention arm reporting diarrhoea in the previous week compared to 31.7% in the control arm. The intervention group had consistently worse anthropometric indicators of undernutrition: mean WHZ [-3.23 versus -3.18], mean height-for-age z-score (HAZ) [-3.29 versus -3.25], worse mean MUAC [117.4 versus 118.7].

**Table 1:**
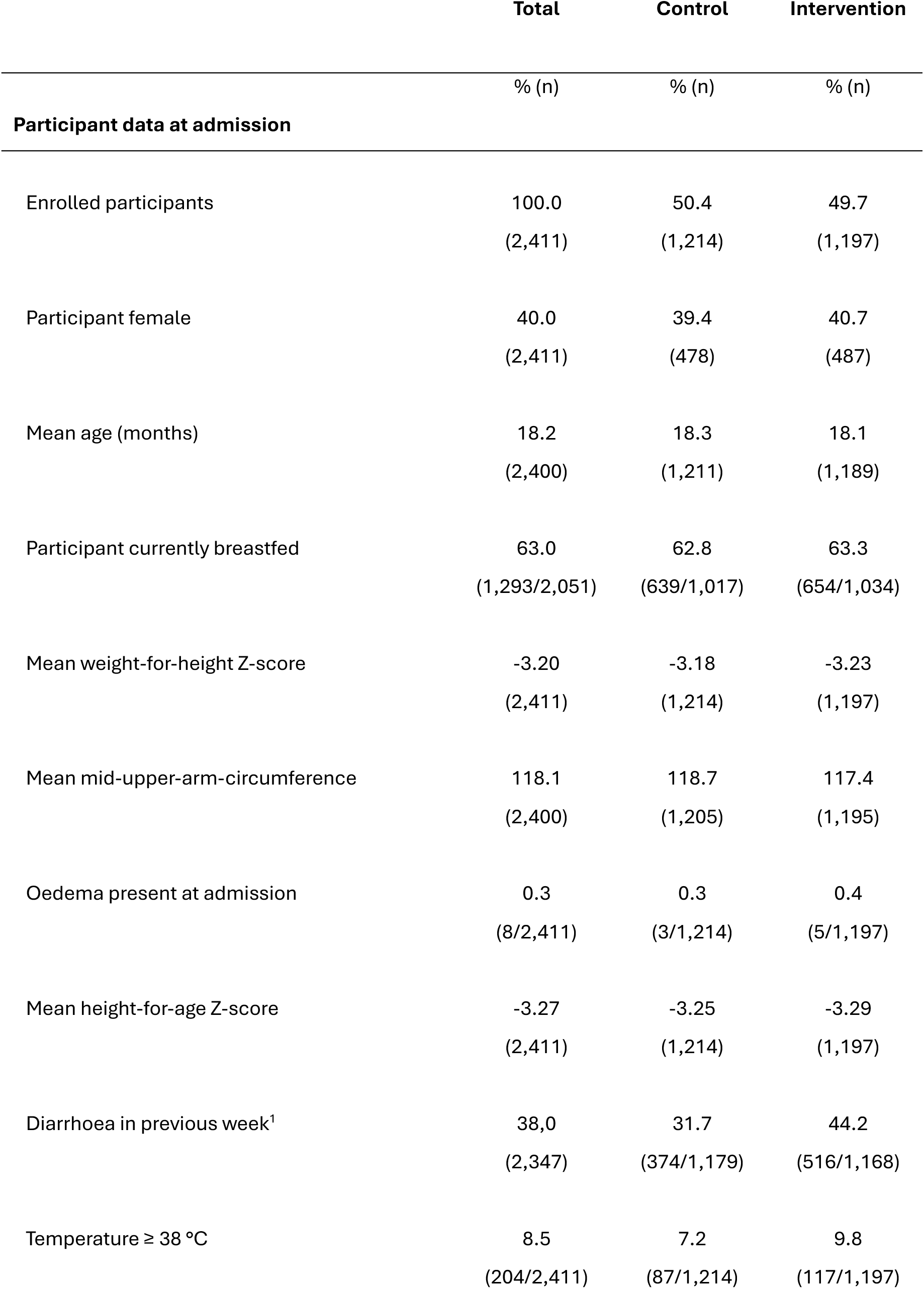
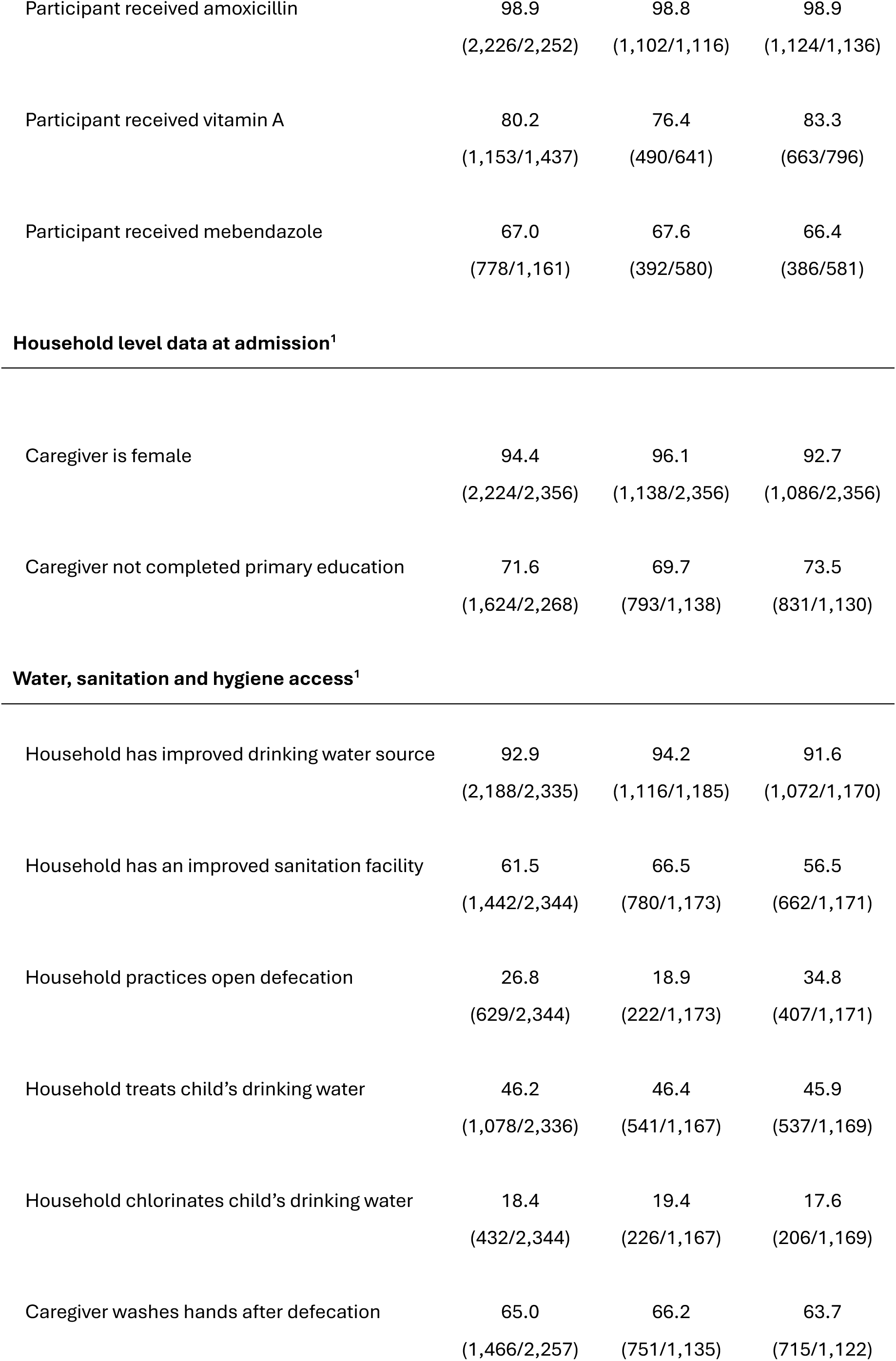
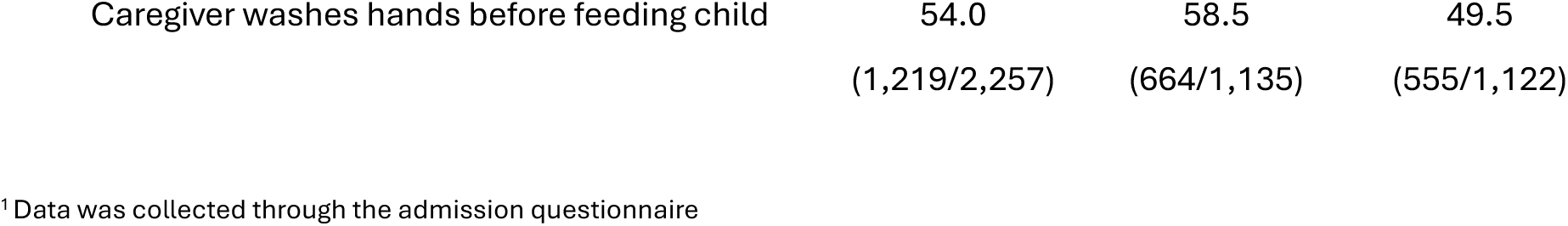
Baseline characteristics in control and intervention groups.

### Water, sanitation and hygiene

Household WASH conditions varied between arms (Table 1) with worse conditions in the intervention arm compared to the control arm for some aspects. A lower proportion of households in the intervention group had access to an improved sanitation facility (56.5%) compared to the control group (66.5%), while a larger proportion reported open defecation (34.8% versus 18.9%, respectively). Reported caregiver handwashing practices were generally lower in the intervention arm versus the control arm but this was most pronounced for handwashing before feeding (49.5% versus 58.5%). Access to an improved drinking water source was greater than 90% in both arms but slightly lower in the intervention arm (91.6% versus 94.2%) and reported chlorination of drinking water was similar across arms (17.6% intervention versus 19.4% control).

### Intervention delivery

In the intervention arm, 96.8% received the WASH kit, while 88.4% received it as intended at admission to CMAM treatment, with the remainder receiving it at a subsequent visit to the health center (Table 2). Less than 1% (5/1214) of participants in the control arm received a WASH kit at admission. Households in both arms were visited by the research team at approximately four weeks post-intervention to assess the presence of the WASH kit as well as reported behaviours around water treatment and handwashing. In 97.4% of the households visited in the intervention arm, the WASH kit container was being used to store drinking water consumed by the participant. The stored drinking water was reported as being treated in 82.6% of households in the intervention arm compared to 8.2% in the control arm. More caregivers in the intervention arm (94.7%) than the control arm (87.0%) reported using soap for handwashing versus using water alone. The proportion of caregivers reporting handwashing after defecation was similar in the two arms but handwashing before feeding was higher in the intervention arm (63.1%) than in the control arm (53.7%).

**Table 2:** Intervention fidelity and response.

|  | Total | Control | Intervention |
| --- | --- | --- | --- |
| Intervention fidelity and response | % (n) | % (n) | % (n) |
| WASH kit provided at admission as planned* | 44.1<br>(1,063/2,411) | 0.4<br>(5/1,214) | 88.4<br>(1,058/1,197) |
| Child drinking water stored in TISA container** | 49.01<br>(820/1,673) | 0.7<br>(6/837) | 97.4<br>(814/836) |
| Stored drinking water treated at household ** | 45.4<br>(761/1,677) | 8.2<br>(69/839) | 82.6<br>(692/838) |
| Caregiver uses soap for handwashing ** | 90.9<br>(1,508/1,659) | 87.0<br>(717/824) | 94.7<br>(791/835) |
| Caregiver washes hands after defecation ** | 79.5<br>(1,254/1,577) | 78.7<br>(612/778) | 80.4<br>(642/799) |
| Caregiver washes hand before feeding child ** | 58.4<br>(921/1,576) | 53.7<br>(417/777) | 63.1<br>(503/799) |

\* The WASH kit was provided on the day of admission to the intervention group as per protocol
\*\*Reported by caregiver at 4-week household visit

### Primary and secondary outcomes

For the primary outcome, ascertained through weight, height and/or MUAC values recorded in health registries, the proportion of children recovering from SAM was similar in the control (39.8%) and intervention groups (41.2%) and we found no evidence for a difference between arms (adjusted odds ratio [aOR] 0.97, 95% confidence interval [CI] 0.66, 1.48) (Table 3). Recovery, as recorded by nurses, was higher in the intervention arm (60.6%) than the control arm (55.3%) but with no evidence of a difference (aOR: 1.08; 95%CIs: 0.72-1.64) (Supplementary Annex 4).

**Table 3:** Adjusted and crude effects for the primary and secondary trial outcomes.

|  | Control |  | Intervention |  | Crude effect |  |  | Adjusted<sup>1</sup> effect |  |  |
| --- | --- | --- | --- | --- | --- | --- | --- | --- | --- | --- |
|  | n/N | % | n/N | % | Odds ratio | 95% CI | p-value | Odds ratio | 95% CI | p-value |
| **Primary outcome** |  |  |  |  |  |  |  |  |  |  |
| <u>Recovery<sup>2</sup></u> | 324/815 | 39.8 | 314/763 | 41.2 | 0.98 | 0.66, 1.46 | 0.93 | 0.97 | 0.66, 1.48 | 0.947 |
| **Secondary outcomes** |  |  |  |  |  |  |  |  |  |  |
| <u>Rate of referral</u> | 16/1214 | 1.32 | 13/1197 | 1.09 | 0.79 | 0.30, 2.07 | 0.634 | 0.75 | 0.29, 1.99 | 0.569 |
| <u>All-cause mortality</u> | 5/1214 | 0.41 | 3/1197 | 0.25 | 0.59 | 0.13, 2.75 | 0.501 | 0.59 | 0.12, 2.90 | 0.512 |
| <u>Diarrhoea prevalence</u> |  |  |  |  |  |  |  |  |  |  |
| Week 0 | 374/1179 | 31.7 | 516/1168 | 44.2 | 1.00 | - | <0.001 | 1.00 |  | <0.001 |
| Week 4 | 266/836 | 31.8 | 246/839 | 29.3 | 0.57 | 0.44, 0.75 |  | 0.57 | 0.43, 0.75 |  |
| Week 8 | 151/754 | 20.0 | 96/769 | 12.5 | 0.36 | 0.26, 0.50 |  | 0.36 | 0.26, 0.50 |  |
| <u>Pathogen detection</u> | 488/782 | 62.40 | 455/724 | 62.85 | 1.16 | 0.82, 1.56 | 0.397 | 1.15 | 0.79, 1.65 | 0.470 |
|  | **Mean** | **SE** | **Mean** | **SE** | **Difference** | **95% CI** | **p-value** | **Difference** | **95% CI** | **p-value** |
| <u>Weight gain, g/kg/d</u> | 4.06<br>(N = 1135) | 0.12 | 4.38<br>(N=1129) | 0.17 | 0.24 | -0.40, 0.88 | 0.466 | 0.18 | -0.46, 0.82 | 0.578 |

<sup>1</sup> Adjusted for age, sex, diarrhoea and HAZ at admission, and household sanitation, water source
<sup>2</sup> As stated in the methods, for recovery, the denominator includes only participants for whom treatment outcome is known at study exit (i.e. participants who recovered, not recovered, referred, and deaths) and excludes those whose status is not known (i.e. those who have defaulted or have been transferred).

Weight gain was greater in the intervention arm (4.38 g/kg/d) than the control arm (4.06 grams per kg per day [g/kg/d]) with no evidence of a difference between arms (difference in means 0.18; 95%CIs: -0.46 - 0.82). Fewer participants were referred in the intervention arm (1.1%) than the control arm (1.3%) with no evidence of a difference between groups (aOR: 0.75; 95% CI: 0.29, 1.99). Five deaths occurred in the control arm [0.4%]) and three in the intervention arm (0.3%) with no evidence of a difference between arms (aOR: 0.59, 95% CI: 0.12, 2.90) (Table 3).

The prevalence of caregiver-reported diarrhoea at eight weeks follow-up was lower in the the intervention group (12.5%) than the control group (20.0%) with evidence of a difference between arms, accounting for between-group differences at admission (aOR: 0.36; 95%CI: 0.26, 0.50) (Table 3). There was also an effect albeit smaller at four weeks post-admission (aOR: 0.57; 95%CI: 0.43 – 0.75).

The enteric pathogen detection prevalence at eight weeks follow-up was very similar in the two arms, with 62.9% and 62.4% of participants positive for at least one of the assessed enteric pathogens in the intervention and control arms respectively (Table 3). There was no evidence of a difference (aOR: 1.15; 95%CI: 0.79, 1.65) in enteric pathogen detection prevalence between arms (Table 3).

## Discussion

In this pragmatic trial, we assessed the effectiveness of integrating drinking water treatment and hand hygiene promotion into the national protocol for outpatient treatment of SAM in Senegal. We sought to assess the effectiveness of such an intervention when delivered at scale and through the existing structure of the health system. The intervention was successfully delivered at scale with almost 90% of participants in the intervention arm receiving the WASH kit on the day of admission into the existing OTP. Furthermore, in household visits approximately one month later, almost all (97%) households were using the designated container for drinking water storage and over 80% were treating their water as recommended with chlorine tablets. However, we found no difference in the proportion of children recovering from SAM in the group receiving the water treatment and hygiene intervention in addition to the standard protocol. There was also no difference in the related outcomes of weight gain during treatment, referral to hospital care nor all-cause mortality. However, diarrhoea was reduced in the intervention group at four weeks and eight weeks of follow-up.

Our trial was designed to assess the effectiveness of integrating this water and hygiene intervention within existing policy and systems and at a scale that would provide evidence for deployment at a national level. The trial was successfully implemented in 86 health centres, over two years of enrolment, and across 28,000 km^2^ which accounts for over 10% of the land mass of Senegal. Given the scale and the challenges encountered during enrolment that included the COVID-19 pandemic, severe flooding, and health system strikes (16), it is notable that the intervention was successfully delivered by the health system with almost 90% of participants receiving the intervention on the day of admission as intended. Furthermore, when assessed at the household level approximately four weeks post-admission almost 100% of households in the intervention group were using the container, and over 80% treating the water in the container as intended. Handwashing with soap – versus water alone – was higher in the intervention group, as was handwashing before feeding the child, which was intended under the intervention. Similarly, a sub-study of the trial that included 445 participant households, found both higher residual chlorine levels and reduced microbial contamination in the intervention group compared to the control group (25). A major question that informed our trial and the underlying theory of change was whether such an intervention could feasibly be integrated within national policy and systems – as opposed to being delivered by external agents at a more limited scale of a project or programme within a discrete area or population as in previous studies. Our results suggest that this approach is feasible, although the costs and benefits would need to be carefully considered in any given setting.

Two previous trials assessed the effectiveness of integrating drinking water treatment and/or improved hand hygiene in CMAM programmes and both reported improved SAM outcomes in the intervention group. The settings for these two trials – the Kanem region of Chad (9) and the Sindh province of Pakistan (8) – are quite different to this study setting. Whilst there is a high burden of acute malnutrition in northern Senegal, it is a stable setting with relatively high health system coverage, and a well-established protocol for CMAM (PECMA) that is administered directly by the government of Senegal. By contrast, the trials in Chad and Pakistan took place in settings with generally lower levels of access to health services where the CMAM programmes were delivered directly by external humanitarian agencies rather than by the Ministry of Health as in our study. Access to safe water and sanitation services in the study populations for these trials appeared worse than in our study, suggesting a higher risk of environmental exposure to enteric pathogens. In the trial in Pakistan, access to improved drinking water and sanitation ranged from 82.3-94.1% and 30-42% respectively across the study arms (8), compared to 92.9% and 61% in our trial. The trial in Chad (9) did not report on water and sanitation services but a separate case-control study (26) by the same team in the same population reported that 46.5% of the population practiced open defecation and 23.9% used unimproved drinking water sources, which was much higher than in our study population. Possibly linked to these underlying factors, admission criteria (WHZ, MUAC, presence of oedema) were consistently worse among participants in these two other trials compared to this one.

Our main finding – that the integration of a water treatment and hygiene promotion intervention into the standard national protocol for outpatient treatment of uncomplicated SAM did not improve recovery outcomes – differs from these two earlier trials with similar interventions (8, 9). Both of these previous studies reported significant increases in the proportion of children recovering from SAM among those receiving an integrated drinking water treatment and/or hygiene. The Chad adopted a similar design – a cRCT with allocation by clinic – to evaluate a similar intervention – the integration of a “WASH kit” into a standard CMAM approach – and found an absolute difference of 10.5% (95% CI: 6.7, 19.8) in the proportion recovering between groups (9). The trial in Pakistan, described as a “site-randomised trial”, evaluated the effectiveness of integrating three different drinking water treatment interventions, chlorine tablets, a double-action flocculant/disinfectant product, and ceramic filters. All three of these water treatment interventions were found to be associated with increased recovery rates compared to a control site receiving standard CMAM. Chlorine tablets, as included in our intervention, were associated with the greatest odds of recovery (aOR: 2.5; 95% CI:1.7, 3.9).

There are three important differences in how recovery was assessed in the previous two trials compared to ours. First, we assessed recovery at eight weeks post-admission compared to 12 weeks in the Chad trial (9) and 17 weeks in the Pakistan trial (8). Eight weeks is the recommended acceptable period for outpatient treatment before referral under the PECMA protocol for Senegal (19). Given that the proportion recovered is cumulative, it is likely that recovery rates in our trial would have been greater if follow-up had been longer, but we cannot know whether this would have resulted in a difference between arms. Second, the criteria for recovery differed in these trials compared to ours. In Chad, if a patient was admitted on WHZ (<-3.0), recovery required the patient to achieve a WHZ of ≥-2.0 compared to ≥ -1.5 in our study. In Pakistan, patients were assessed for recovery on MUAC (≥125mm) only. Third, the nature, dosage and intensity of these interventions differed to ours with the TISA trial providing fewer products but placing greater emphasis on sensitization, including home visits, compared to the other two trials.

We found a large effect on the prevalence of caregiver-reported diarrhoea at both eight weeks (aOR: 0.36; 95% CI: 0.26, 0.50) and four weeks post-admission (aOR: 0.57; 95% CI: 0.44, 0.75). This effect estimate accounts for the higher prevalence of diarrhoea at admission in the intervention group compared to the control group. By comparison, the trial in Chad found no effect on child diarrhoea but the trial in Pakistan found that chlorine tablets – the same type as used in our trial – but not the other water treatment interventions (double-action disinfectant/flocculant or ceramic filter) were associated with a lower risk of diarrhoea. Notably, the prevalence of diarrhoea at admission was lower in Chad than in Pakistan or in our study.

Our trial included enteric pathogen detection as a secondary outcome but found no effect on the prevalence of detecting one or more pathogens in participants’ stool. There are several advantages of using stool-based enteric pathogen detection via multiplex methods in trials of WASH interventions (27). Notably they provide an objective measure of exposure to a given pathogen that lies on the causal pathway between environmental hazards and disease outcomes. In our trial, we incorporated these methods to enhance our understanding of if - and how - the intervention modified participants’ exposure to environmental hazards. A recent meta-analysis found that WASH interventions were only associated with small reductions in enteric pathogen detection in the environment but only basic sanitation interventions were included (28), and not water treatment or hand hygiene interventions such as were evaluated in our trial. A separate individual participant data analysis assessed the association between enteric pathogen detection in the environment (soil, children’s hands, and stored drinking water) and subsequent detection in children, childhood diarrhoea and growth faltering (29). Interestingly, environmental detection was associated with increased risk of detection in children and lower HAZ but not diarrhoea (29). There have been few WASH trials incorporating enteric pathogen detection as outcomes and, to our knowledge, this is the first WASH trial to do so in the context of SAM treatment and recovery. Whilst not among SAM outpatients, one trial of a water treatment technology that included enteric pathogen detection as an outcome reported no effect in a community setting (30). Future analysis of our data at the individual pathogen level will allow us to assess the effect of the intervention on specific pathogens and to explore the relationships between environmental factors, enteric pathogen detection, diarrhoea and nutritional status.

## Limitations

Despite the random allocation of the treatment by cluster, there appeared to be imbalance between arms for certain covariates. At baseline, the intervention group had a lower proportion of participants with access to an improved sanitation facility (56.5% versus 66.5%) and a lower proportion of caregivers reported washing their hands before feeding their child (49.5% versus 58.5%) compared to the control group. There was no difference however for the proportion with access to an improved drinking water source or who reported treating their drinking water. The one-week period prevalence of diarrhoea was higher in the intervention group compared to the control group at admission (31.7% versus 44.2%). We compared pathogen detection only at endline meaning that we could not adjust for differences at baseline between groups which may well have been present given the observed imbalance for diarrhoea. Whilst our analyses were adjusted for these covariates on the basis of this observed imbalance, we took a decision to not use a balancing method in our randomization on the basis of the large number of clusters enrolled but those designing future trials might consider this. The previous trial in Chad randomised in stratified pairs according to monthly admission numbers to balance enrolment across arms but still had significant imbalance for diarrhoea on admission. A recent analysis has demonstrated that random allocation within geographically matched pairs improved statistically efficiency across all considered outcomes, including outcomes relevant to our trial (31). Such an approach would have been feasible for our trial and may have yielded greater balance and statistical efficiency.

We sought to assess the real-world effectiveness – and feasibility – of this intervention when integrated into existing health policy and systems. Such trials can be termed “pragmatic” and are generally motivated by concerns as to the value for policy of trial results where efficacy of interventions has been optimized (32). Our trial revealed underlying challenges that may be less apparent in more controlled trial settings or indeed in humanitarian settings where populations are bound to limited geographies due to security and/or availability of shelter and services. The observed rate of defaulting was very high but similar to the official figures. This likely reflects challenges for households in balancing the benefits of continuing treatment versus the financial and time costs for accessing treatment when facilities are distant and the relative cost of transport high. A further contributing factor is the “transhumance” - nomadic pastoralism whereby families move with their animals to seek pasture during dry seasons - which makes accessing and sustaining treatment challenging (33). Our trial was designed to estimate the effect of integrating this intervention within the existing health systems and, as such, our results account for these factors by design and provide evidence that can inform investment decisions in this setting. The high rate of defaulting suggests that interventions or targeted approaches to tackle the high rate of defaulting before recovery may yield improved outcomes.

## Conclusions

In this setting of northern Senegal, the integration of household water treatment and hygiene promotion to the standard treatment protocol did not improve SAM recovery nor the related outcomes of weight gain and referral. There was no reduction in stool-based enteric pathogen detection, but the intervention did reduce diarrhoea. Our results suggest that, in this setting, the addition of a WASH kit to the standard protocol would not improve SAM outcomes but could potentially reduce the burden of diarrhoea among this vulnerable group.

## Data Availability

All data produced in the present study are available upon reasonable request to the authors

