## Supplemental Information for "Effect of household water treatment and hygiene promotion integration on outpatient treatment for severe acute malnutrition in Senegal: the TISA cluster-randomised controlled trial"

**Supplementary Information**:

SI Figure 1: Monthly enrolment rate for all participants and by control and intervention arm

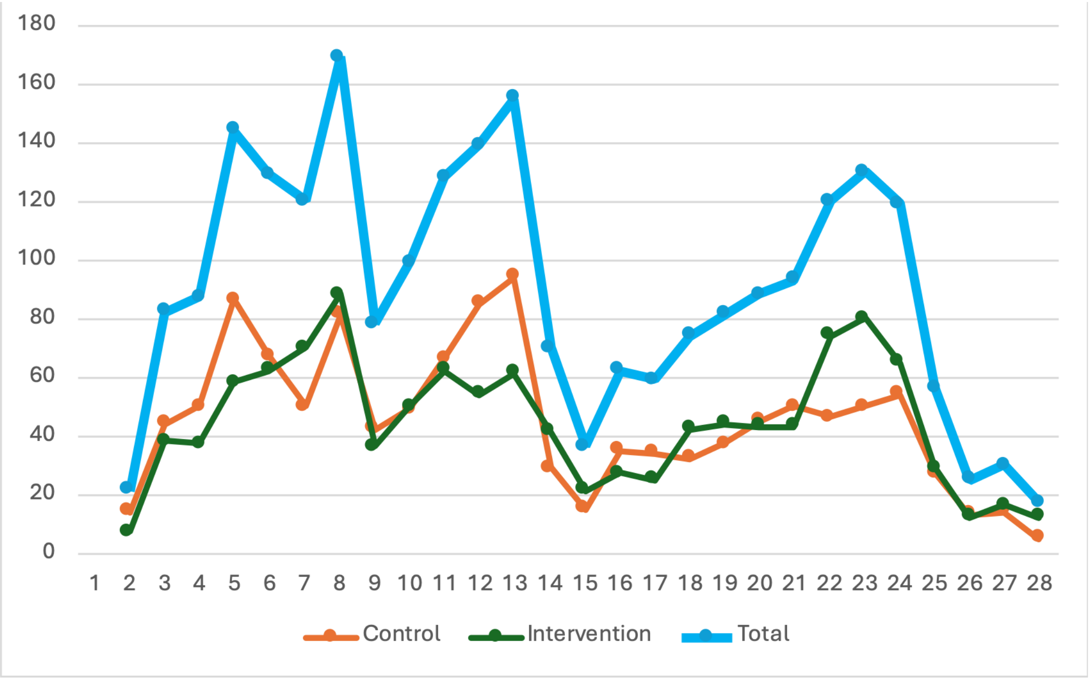

### SI Table 2: Outpatient treatment outcomes at study exit

#### *Table 2.1 Per study protocol treatment outcomes by arm*

Under the study protocol, participants were considered recovered if the participant achieved two consecutive measures at weekly health centre visits with WHZ ≥ -1.5, if admitted based on WHZ, and/or MUAC ≥ 125 mm, if admitted based on MUAC, and no oedema.

| **Study exit (Protocol)** | **Total** | | **Control** | | **Intervention** | |
|---|---|---|---|---|---|---|
| | n | % | n | % | n | % |
| **Defaulted** | 777 | 32.2 | 366 | 30.2 | 411 | 34.3 |
| **Recovered** | 638 | 26.5 | 324 | 26.7 | 314 | 26.2 |
| **Non-recovered** | 931 | 38.6 | 484 | 39.9 | 447 | 37.3 |
| **Referred** | 29 | 1.2 | 16 | 1.3 | 13 | 1.1 |
| **Transferred** | 28 | 1.2 | 19 | 1.6 | 9 | 0.8 |
| **Death** | 8 | 0.3 | 5 | 0.4 | 3 | 0.3 |
| **Total** | 2,411 | 100.0 | 1,214 | 100.0 | 1,197 | 100.0 |

#### *Table 2.2 Per health registry treatment outcomes by arm*

This table presents treatment outcomes as recorded in the health registry by nurses. Participants were often recorded as having recovered when meeting recovery criteria for a single week and not for two consecutive weeks as required under our study protocol.

| **Study exit (Registry)** | **Total** | | **Control** | | **Intervention** | |
|---|---|---|---|---|---|---|
| | n | % | n | % | n | % |
| **Defaulted** | 591 | 24.5 | 303 | 25,0 | 288 | 24.1 |
| **Recovered** | 1,038 | 43.1 | 493 | 40.6 | 545 | 45.5 |
| **Non-recovered** | 717 | 29.7 | 378 | 31.0 | 339 | 28.3 |
| **Referred** | 29 | 1.2 | 16 | 1.3 | 13 | 1.1 |
| **Transferred** | 28 | 1.2 | 19 | 1.6 | 9 | 0.8 |
| **Death** | 8 | 0.3 | 5 | 0.4 | 3 | 0.3 |
| **Total** | 2,411 | 100.0 | 1,214 | 100.0 | 1,197 | 100.0 |

SI Figure 2: One week prevalence of diarrhoea at 0, 4 and 8 weeks of follow-up by trial arm

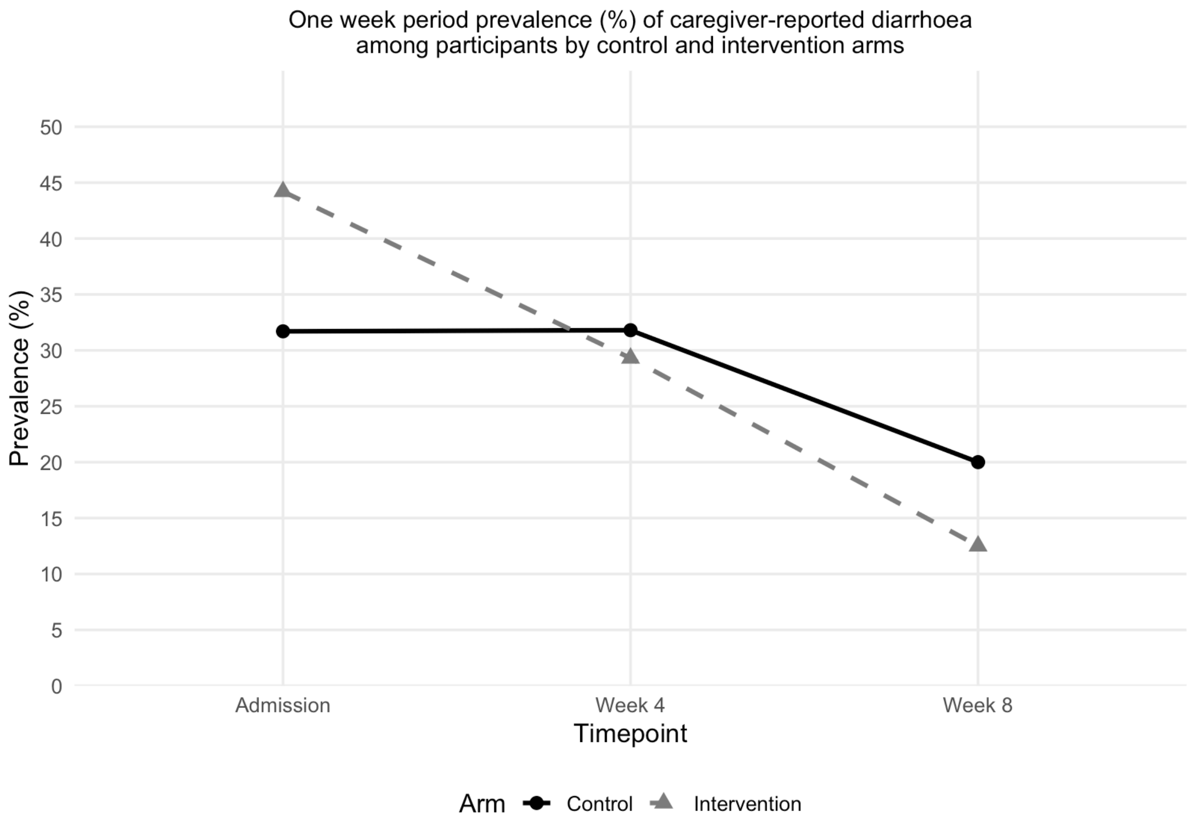

### SI Table 3: Adjusted and crude effects of intervention on health registry recorded recovery

| | Control | | Intervention | | Crude effect | | | Adjusted[1] effect | | |
|---|---|---|---|---|---|---|---|---|---|---|
| | n/N | % | n/N | % | Odds ratio | 95% CI | p-value | Odds ratio | 95% CI | p-value |
| **Nurse recorded recovery** | 493/892 | 55.3 | 545/900 | 60.6 | 1.11 | 0.73, 1.69 | 0.63 | 1.08 | 0.72, 1.64 | 0.700 |

[1] Adjusted for age, sex, diarrhoea at admission, sanitation, water source and HAZ at admission

### SI Table 4: TaqMan Array Card molecular gene targets and sequences

| Category | Pathogen | Target | reference | Primers & probes sequences (labelled with FAM (6-carboxyfluorescein) at 5' and MGB at 3') |
|---|---|---|---|---|
| **Virus** | Adenovirus F (40/41) | fiber gene | (1) | F: AACTTTCTCTCTTAATAGACGCC; R: AGGGGGCTAGAAAACAAAA<br>Probe: CTGACACGGGCACTCT |
| | Astrovirus | Capsid | (2) | F: CAGTTGCTTGCTGCGTTCA; R: CTTGCTAGCCATCACACTTCT<br>Probe: CACAGAAGAGCAACTCCATCGC |
| | Norovirus GI | ORF1-2 | (1) | F: CGYTGGATGCGNTTYCATGA; R: CTTAGACGCCATCATCATTYAC<br>Probe: TGGACAGGAGATCGC |
| | Norovirus GII | ORF1-2 | (2,3) | F: CARGARBCNATGTTYAGRTGGATGAG; R: TCGACGCCATCTTCATTCACA<br>Probe: TGGGAGGGCGATCGCAATCT |
| | Rotavirus | NSP3 | (2,4) | F: ACCATCTWCACRTRACCCTCTATGAG; R: GGTCACATAACGCCCCTATAGC<br>Probe: AGTTAAAAGCTAACACTGTCAAA |
| | Sapovirus | RdRp | (2) | Fw1: GAYCASGCTCTCGCYACCTAC; Fw2: TTGGCCCTCGCCACCTAC;<br>R: CCCTCCATYTCAAACACTA<br>Probe: CCRCCTATRAACCA |
| **Bacteria** | Aeromonas | aerolysin | (2) | F: TYCGYTACCAGTGGGACAAG; R: CCRGCAAACTGGCTCTCG<br>Probe: CAGTTCCAGTCCCACCACTT |
| | Campylobacter jejuni/coli | cadF | (2,5) | F: CTGCTAAACCATAGAAATAAAATTTCTCAC; R: CTTTGAAGGTAATTTAGATATGGATAATCG<br>Probe: CATTTTGACGATTTTTGGCTTGA |
| | C. difficile | tcdB | (2) | F: GGTATTACCTAATGCTCCAAATAG; R: TTTGTGCCATCATTTTCTAAGC<br>Probe: CCTGGTGTCCATCCTGTTTC |
| | EAEC | aaiC | (2,6) | F: ATTGTCCTCAGGCATTTCAC; R: ACGACACCCCTGATAAACAA<br>Probe: TAGTGCATACTCATCATTTAAG |
| | EAEC | aatA | (2,6) | F: CTGGCGAAAGACTGTATCAT; R: TTTTGCTTCATAAGCCGATAGA<br>Probe: TGGTTCTCATCTATTACAGACAGC |
| | EAEC | aggR | (1,7) | F: GCAATCAGATTAARCAGCGATACA; R: TTCGGACAACTRCAAGCATC<br>Probe: AAGACGCCTAAAGGATGCCC |
| | STEC | stx1 | (2) | F: ACTTCTCGACTGCAAAGACGTATG; R: ACAAATTATCCCCTGWGCCACTATC<br>Probe: CTCTGCAATAGGTACTCCA |
| | STEC | stx2 | (2,8) | F: CCACATCGGTGTCTGTTATTAACC; R: GGTCAAAACGCGCCTGATAG<br>Probe: TTGCTGTGGATATACGAGG |
| | EPEC | Eae | (2) | F: CATTGATCAGGATTTTTCTGGTGATA; R: CTCATGCGGAAATAGCCGTTA<br>Probe: ATACTGGCGAGACTATTTCAA |
| | EPEC | bfpA | (1,2) | F: TGGTGCTTGCGCTTGCT; R: CGTTGCGCTCATTACTTCTG<br>Probe: CAGTCTGCGTCTGATTCCAA |
| | ETEC | LT | (2,8) | F: TTCCCACCGGATCACCAA; R: CAACCTTGTGGTGCATGATGA<br>Probe: CTTGGAGAGAAGAACCCT |
| | ETEC | STh | (2) | F: GCTAAACCAGYAGRGTCTTCAAAA; R: CCCGGTACARGCAGGATTACAACA<br>Probe: TGGTCCTGAAAGCATGAA |
| | ETEC | STp | (2) | F: TGAATCACTTGACTCTTCAAAA; R: GGCAGGATTACAACAAAGTT<br>Probe: TGAACAACACATTTTACTGCT |
| | E. coli O157 | rfbE | (1,9( | F: TTTCACACTTATTGGATGGTCTCAA; R: CGATGAGTTTATCTGCAAGGTGAT |

| | | | | |
|---|---|---|---|---|
| | | | | Probe: CTCTCTTTCCTCTGCGGTCCT |
| | Helicobacter pylori | ureC | (1) | F: GACACCAGAAAAAGCGGCTA; R: AGCGCATGTCTTCGGTTAAA<br>Probe: TCACTAAAGCGTTTTCTACC |
| | Plesiomonas shigelloides | gyrB | (1) | F: CCGCCGTGAAGGCAAAG; R: GCTACCGGCTCACCCAGAT<br>Probe: CACACCCAAGAATAC |
| | Salmonella enterica | ttr | (1,10) | F: CTCACCAGGAGATTACAACATGG; R: AGCTCAGACCAAAAGTGACCATC<br>Probe: CACCGACGGCGAGACCGACTTT |
| | Salmonella enterica Typhi | STY0201 | (1,11) | F: CGCGAAGTCAGAGTCGACATAG; R: AAGACCTCAACGCCGATCAC<br>Probe: CAGCCTGCTCCAGAACA |
| | Shigella/EIEC | ipaH | (1,12) | F: CCTTTTCCGCGTTCCTTGA; R: CGGAATCCGGAGGTATTGC<br>Probe: CGCCTTTCCGATACCGTCTCTGCA |
| | Shigella flexneri | Putative periplasmic Protein* | (13) | F: TGGGTGCATCCTGACCTGT; R: GACAAACAATAACGAGCTACCGAT<br>Probe: ACCACGGAATAATCCCGCAG |
| | Shigella flexneri | O-antigen** | (13) | F: CTCCTATCCGTGATTATAGTGCA; R: GCACACACAACTCACTGTATTT<br>Probe: TCCTTCTCACGATTAAAATC |
| | Shigella flexneri | Type 3 restriction Enzyme** | (13) | F: CTTTCAACGCACGAATATCAAC; R: GAACCTGATCCAGACGGAGA<br>Probe: TTCTTCAGAACCGGGTTTTG |
| | Shigella sonnei | Putative methylase | (13) | F: TGCCGCTAAAATCCTTCTGT; R: GCGTACGACGAAAGGAAAAA<br>Probe: GAAGTTATTGATTCCGCCC |
| | Vibrio cholerae | hlyA | (1) | F: ATCGTCAGTTTGGAGCCAGT; R: TCGATGCGTTAAACACGAAG<br>Probe: ACCGATGCGATTGCCCAA |
| | Vibrio cholerae | ctxA | (14) | F: GCATAGAGCTTGGAGGGAAGAG; R: CATCGATGATCTTGGAGCATTC<br>Probe: CATCATGCACCGCCG |
| | Yersinia enterocolitica | lytA | (1,15) | F: TGATTCACCAGCAGCAATAC; R: GGCATCATGAAAGGCGG<br>Probe: TGTCGGTTTCTCCTTCCAGG |
| **Protozoa** | Cryptosporidium spp. | 18S rRNA | (1) | F: GGGTTGTATTTATTAGATAAAGAACCA; R: AGGCCAATACCCTACCGTCT<br>Probe: TGACATATCATTCAAGTTTCTGAC |
| | Cyclospora cayetanensis | 18S | (1) | F: AAAAGCTCGTAGTTGGATTTCTG; R: AACACCAACGCACGCAGC<br>P: AAGGCCGGATGACCACGA |
| | Giardia spp. | 18S rRNA | (2,16) | F: GACGGCTCAGGACAACGGTT; R: TTGCCAGCGGTGTCCG<br>Probe: CCCGCGGCGGTCCCTGCTAG |
| | E. histolytica | 18S rRNA | (2,16) | F: ATTGTCGTGGCATCCTAACTCA; R: GCGGACGGCTCATTATAACA,<br>Probe: TCATTGAATGAATTGGCCATTT |
| **Helminth** | Ascaris lumbricoides | ITS1 | (1) | F: GCCACATAGTAAATTGCACACAAAT; R: GCCTTTCTAACAAGCCCAACAT<br>Probe: TTGGCGGACAATTGCATGCGAT |
| | Trichuris trichiura | 18S rRNA | (2) | F: TTGAAACGACTTGCTCATCAACTT; R: CTGATTCTCCGTTAACCGTTGTC<br>Probe: CGATGGTACGCTACGTGCTTACCATGG |
| | Ancylostoma duodenale | ITS2 | (1,17) | F: GAATGACAGCAAACTCGTTGTTG; R: ATACTAGCCACTGCCGAAACGT<br>Probe: ATCGTTTACCGACTTTAG |
| | Necator americanus | ITS2 | (1,17) | F: CTGTTTGTCGAACGGTACTTGC; R: ATAACAGCGTGCACATGTTGC<br>Probe: CTGTACTACGCATTGTATAC |
| | Strongyloides stercoralis | dispersed repetitive sequence | (1,18) | F: TCCAGAAAAGTCTTCACTCTCCAG; R: TGCGTTAGAATTTAGATATTATTGTTGCT<br>Probe: TCAGCTCCAGTTGAACAACAGCCTCCAA |

| | | | | |
|---|---|---|---|---|
| | Schistosoma spp. | ITS | (1,19) | F: GGTCTAGATGACTTGATYGAGATGCT; R: TCCCGAGCGYGTATAATGTCATTA<br>P: TGGGTTGTGCTCGAGTCGTGGC |
| **Control/ RNA virus** | MS2 | MS2g1 | (2,22) | F: TGGCACTACCCCTCTCCGTATTCAC; R: GTACGGGCGACCCCACGATGAC<br>Probe: CACATCGATAGATCAAGGTGCCTACAAGC |
| **Control/ DNA virus** | PhHV | gB | (2) | F: GGGCGAATCACAGATTGAATC; R: GCGGTTCCAAACGTACCAA<br>Probe: TATGTGTCCGCCACCATCT |
| **Control/ 16S rRNA** | 16S | 16S | (1,23) | F: TGCAAGTCGAACGAAGCACTTTA; R: GCAGGTTACCCACGCGTTAC<br>Probe: CGCCACTCAGTCACAAA |
| **Control/ 18S rRNA** | 18S† | 18S | N/A | Manufacturer's control |
